# Patient preferences and priorities for the design of an acute kidney injury prevention trial: Findings from a consensus workshop

**DOI:** 10.1101/2024.03.04.24303721

**Authors:** Meghan J. Elliott, Kirsten M. Fiest, Shannan Love, Dale Birdsell, Maureena Loth, Heather Dumka, Benny Rana, Nusrat Shommu, Eleanor Benterud, Sarah Gil, Dilaram Acharya, Tyrone G. Harrison, Neesh Pannu, Matthew T. James

## Abstract

**Introduction:** High-quality clinical trials are needed to establish the safety, efficacy, and real-world use of potential therapies for acute kidney injury (AKI) prevention. In this consensus workshop, we identified patient and caregiver priorities for recruitment, intervention delivery, and outcomes of a clinical trial of cilastatin to prevent nephrotoxic AKI.

**Methods:** We included adults with lived experience of AKI, chronic kidney disease, or risk factors for AKI (e.g., critical care hospitalization), and their caregivers. Using a modified nominal group technique approach, we conducted a series of hybrid in-person/virtual discussions covering 3 clinical trial topic areas: (1) consent and recruitment; (2) intervention delivery; and (3) trial outcomes. Participants voted on their top preferences in each topic area, and discussion transcripts were analyzed inductively using conventional content analysis.

**Results:** Thirteen individuals (11 patients, 2 caregivers) participated in the workshop. For consent and recruitment, participants prioritized technology enabled pre-screening and involvement of family members in the consent process. For intervention delivery, participants prioritized measures to facilitate intervention administration and return visits. For trial outcomes, participants identified kidney-related and other clinical outcomes (e.g., AKI, chronic kidney disease, cardiovascular events) as top priorities. Analysis of transcripts provided insight into care team and family involvement in trial-related decisions, implications of allocation to a placebo arm, and impact of participants’ experiences of AKI and critical illness.

**Conclusion:** Findings from our workshop will directly inform development of a clinical trial protocol of cilastatin for nephrotoxic AKI prevention and can assist others in patient-centered approaches to AKI trial design.

## INTRODUCTION

People living with chronic diseases frequently require hospitalization, where they are exposed to a variety of medications.^1–4^ Some of these medications, such as chemotherapeutics for treatment of cancer, immunosuppression for organ transplantation or autoimmune conditions, antibiotics for infections, and contrast dyes for imaging procedures, can cause acute kidney injury (AKI) in up to 25% of patients, especially when used in combination.^5–7^ The consequences of AKI include poor patient outcomes such as prolonged hospitalization, chronic kidney disease (CKD) and kidney failure, cardiovascular events, and death,^6, 8, 9^ as well as health system burden related to high acute and long-term chronic disease care needs.^10^ However, beyond avoidance of potentially implicated medications, which is often neither safe nor feasible for hospitalized patients, no pharmacologic therapies are currently available for prevention of nephrotoxic AKI.^5^

Uptake of nephrotoxins in the proximal tubules of the kidneys is a major contributor to the pathogenesis of AKI.^11–14^ A small molecule called cilastatin can prevent tubular drug uptake and kidney injury through its inhibitory action on two proteins (dipeptidase-1 and megalin).^15–17^ While cilastatin prevents kidney injury in cell culture and animal models of nephrotoxic AKI, it has been only indirectly tested in human trials using a combined formulation approved for clinical use – i.e., imipenem-cilastatin, where cilastatin prevents tubular degradation and extends the action of the antibiotic, imipenem.^18^ In a recent systematic review, pooled results from 10 studies showed lower risks of AKI and better kidney function among patients treated with imipenem-cilastatin compared to inactive or active controls.^19^ Given the lack of alternatives and evidence suggesting fewer adverse events with cilastatin alone than with imipenem-cilastatin formulations already approved for clinical use,^19^ a well-designed and adequately powered trial is needed to establish the efficacy of cilastatin alone for this indication.

Over the past two decades, international initiatives have increasingly integrated the perspectives of patients, or people with lived experience of a health condition, into research activities.^20^ Though patients can meaningfully contribute their expertise as users of the health system at any stage of research, their engagement from the beginning to develop protocols can enhance research feasibility and relevance.^21^ Evidence for involving patients in the co-design of clinical trials includes several benefits, ranging from more patient-centered recruitment practices, informational materials, and outcome selection, to improved experiences for research participants and greater adherence to the trial intervention.^22–24^ Given the individual and health system burden of nephrotoxic AKI and demand for novel therapeutic agents for AKI prevention, integrating patient preferences into the design of an interventional AKI clinical trial is critical for supporting rigorous, pragmatic, and patient-centered research. Therefore, we undertook a consensus-based workshop with people with lived experience of AKI or risk factors for AKI to identify preferences and priorities related to recruitment, intervention delivery, and outcomes for a clinical trial of cilastatin to prevent nephrotoxic AKI.

## METHODS

### Study design and setting

We held a half-day hybrid in-person/virtual workshop in December 2023 at the University of Calgary. Two thirds of participants attended the workshop in person with the remaining participants attending virtually using the Zoom™ platform. We used a modified nominal group technique (NGT),^25^ an accepted consensus building approach, to generate and prioritize preferences related to the design of a clinical trial of cilastatin for nephrotoxic AKI prevention among people with lived experience of AKI or risk factors for AKI. During the workshop, three vignettes (i.e., clinical scenarios involving fictional patients and caregivers) were used to help focus and guide discussions related to three key aspects of clinical trial design: recruitment and consent, intervention delivery, and outcomes (Supplementary Table S1). This study was approved by the Conjoint Health Research Ethics Board at the University of Calgary (REB23-1564).

### Participants and Recruitment

We recruited 13 adult participants who were comfortable communicating in English and who had either experienced or cared for someone with AKI, CKD, and/or risk factors for nephrotoxic AKI. We purposively sampled participants from nephrology and critical care patient advisory groups in Alberta, Canada and among participants from related research expressing interest in being contacted about future studies. Research team members distributed email invitations to potential participants and responded to those indicating interest with additional information about the workshop.

Participants were provided with packages by email one week before the workshop. These packages included a summary of the topic area, workshop agenda, three vignettes (Supplementary Table S1), a consent form, and instructions for parking, lunch, and use of the virtual platform, if required. In the topic summary, definitions and examples of areas for discussion (i.e., recruitment/consent, intervention delivery, and outcomes) were provided. We asked participants to review the vignettes in advance of the workshop and reflect on how a clinical trial in AKI could be designed with the example patients’ and caregivers’ needs in mind. Research team members were available by phone or email to answer questions prior to the workshop and to troubleshoot issues in real time for virtual attendees. All participants provided written informed consent prior to workshop commencement.

### Data collection

An overview of the workshop phases and flow is provided in Figure 1. First, one facilitator (MJ) welcomed participants, explained the purpose of the workshop, and provided a program overview. One virtual and two in-person groups were established, each with 4-5 participants as well as a facilitator (MJ, MJE, KF) and note-taker (EB, SL, BR) with advanced training in qualitative and workshop methodology. Each group participated in three separate small-group discussions for each topic area of trial design – 1) consent and recruitment, 2) intervention delivery, and 3) outcomes. Experienced facilitators led the small-group discussions using a topic guide (Supplementary Table S2) and one of the vignettes to guide the conversation (Supplementary Table S1). Facilitators ensured participants had equal opportunity to contribute by directly inviting them to share their thoughts and views, redirecting the flow of the conversation, and refocusing the discussion around the vignette when required. Following each small group, a facilitator or group participant summarized key points from their group’s discussion for the larger group. Before the final prioritization exercise, research team members met to consolidate and categorize preferences within each topic area.

**Figure 1.**
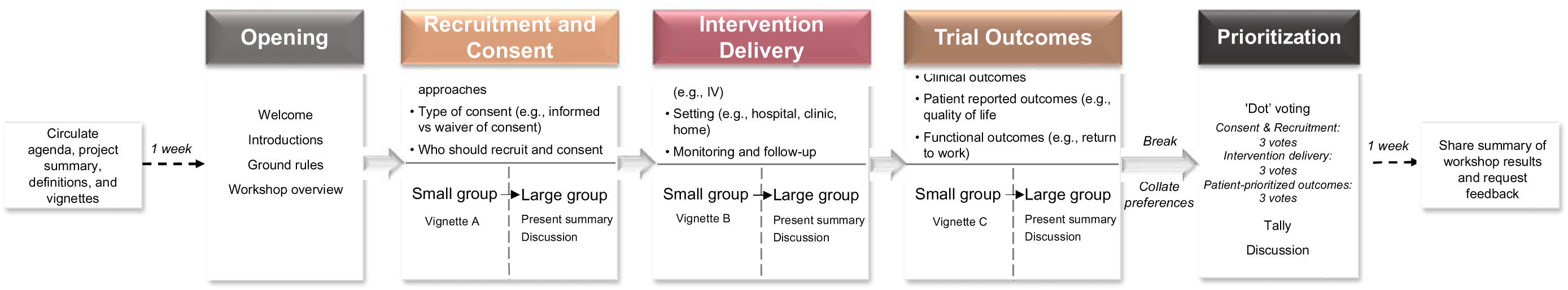
Overview of phases and flow of the consensus workshop.

Using cumulative dot voting,^26, 27^ participants were asked to vote on three preferences that they considered most important under each of the consent and recruitment, intervention delivery, and outcomes topic areas (for a total of 9 dots per participant). Participants voted by either placing a physical dot beside their choice (in person) or by selecting their preferred options using the polling feature on Zoom™ (virtual). All discussions were audio-recorded using a handheld recording device and transcribed by an experienced transcriptionist. One week after the workshop, participants were invited by email to provide their feedback on the workshop format and processes in an evaluation survey (Supplementary Table S3).

### Data analysis

We summarized demographic data provided by participants descriptively. Preferences were ranked in each topic area by tallying the total number of votes and ranking results as high (≤7 votes), medium (3-6 votes), or low (<3 votes) priority. Priority categories were determined based on the number of workshop participants, available selections within each topic area, and results of other similar consensus-based exercises.^28, 29^ Results from the post-workshop evaluation were summarized descriptively (Supplementary Figure S1).

Transcripts from the small and large group discussions were reviewed and inductively analyzed to elaborate on the prioritization results and additional insights raised during group discussions. Using conventional content analysis,^30^ three research team members (MJE, SL, BR) reviewed the transcripts independently and discussed them as a group to generate a list of relevant codes representing distinct ideas. These codes were then sorted into categories, or key concepts, within each of the three topic areas. The key concepts were refined further through discussion among the broader research team and review of handwritten field notes taken during the workshop that captured additional non-verbal cues and group dynamics. We ensured methodological rigour through our transparent and reflexive approach to data collection and analysis, systematic application of consensus-based methods with experienced facilitators, researcher and data triangulation, and provision of rich descriptions to support our findings.^31^

### Patient Engagement

Two patient partners (DB, ML) with lived experience of AKI and/or CKD were part of the core research team supporting development of the cilastatin trial protocol. Both collaborated on the design, conduct, interpretation, and reporting of this project and participated in the workshop. Another patient partner (HD) was the co-lead of the Nephrology Research Group’s Patient and Community Partnership at the University of Calgary and helped develop and coordinate the consensus workshop. Patient partners reviewed final outputs and contributed to manuscript preparation. We shared a graphical summary of the prioritization results and thematic findings with all workshop participants one month after the workshop and invited them to provide feedback, offer alternative interpretations, and request clarification. We have reported this work in accordance with the Guidance for Reporting Involvement of Patients and Public (GRIPP2; Supplementary Table S4).^32^

## RESULTS

Thirteen people participated in the workshop, including four with prior AKI, seven with CKD, six with conditions putting them at risk of AKI (e.g., cardiovascular disease, diabetes, nephrotoxic medication exposure), and two with experience of caregiving for a person with AKI or CKD (Table 1). Seven participants (54%) identified as women, seven (54%) were greater than 65 years of age, and 7 (54%) were retired. Most participants (69%) resided in an urban location. Reduced kidney function (i.e., eGFR of 30-60 mL/min/1.73m^2^) at the time of the workshop was reported by seven (54%) participants, either among themselves or the corresponding patient for caregiver participants; two (15%) patients had received a prior kidney transplant. In the following sections, we summarize results from the prioritization exercise and key concepts arising from small- and large-group discussions in relation to identified priorities (Tables 2 and 3).

**Table 1.** Demographic and clinical characteristics of participants (n=13)

| **Characteristic** | **n (%)** |
| --- | --- |
| **Condition\*** |  |
| Person with previous acute kidney injury (AKI) | 4 (31) |
| *Cause of AKI* |  |
| Critical illness (e.g., sepsis, critical care stay) | 3 |
| Nephrotoxic medication exposure | 1 |
| Person with chronic kidney disease (CKD) | 7 (54) |
| *Cause of CKD* |  |
| Nephrotoxic medication exposure | 2 |
| Prior AKI | 1 |
| Glomerulonephritis | 1 |
| Diabetes | 1 |
| Other (e.g., reflux, polycystic kidney disease) | 2 |
| Person with condition putting them at risk of AKI | 6 (46) |
| *Risk factor\** |  |
| Hypertension | 3 |
| Hospitalization with critical illness | 2 |
| Cardiovascular disease | 1 |
| Diabetes | 1 |
| Other (e.g., nephrotoxic medication exposure) | 1 |
| Caregiver of a person with AKI or CKD | 2 (15) |
| **Place of residence** |  |
| Calgary | 9 (69) |
| Edmonton and northern Alberta | 2 (15) |
| Prefer not to answer | 2 (15) |
| **Age (years)** |  |
| 45 or under | 2 (15) |
| 46-55 | 1 (8) |
| 56-65 | 3 (23) |
| 66-75 | 4 (31) |
| Over 75 | 3 (23) |
| **Education** |  |
| High school | 2 (15) |
| College or trade school | 2 (15) |
| University degree | 5 (8) |
| Professional or graduate degree | 3 (23) |
| Prefer not to answer | 1 (8) |
| **Employment status\*** |  |
| Retired | 7 (54) |
| Part-time or casual | 2 (15) |
| Full-time | 1 (8) |
| Other (e.g., home duties) | 1 (8) |
| Prefer not to answer | 2 (15) |
| **Gender Identity** |  |
| Woman | 7 (54) |
| Man | 6 (46) |
| **Languages spoken\*** |  |
| English | 13 (100) |
| Other (e.g., Dene, Italian) | 3 (23) |
| **Self Reported Race / Ethnicity** |  |
| White | 11 (85) |
| Indigenous | 1 (8) |
| Black African | 1 (8) |
| **Marital status** |  |
| Married | 8 (62) |
| Single | 2 (15) |
| Divorced | 1 (8) |
| Widowed | 1 (8) |
| Prefer not to answer | 1 (8) |
| **Current kidney function**<sup>†</sup> |  |
| eGFR greater than 60 mL/min/1.73 m<sup>2</sup> | 3 (23) |
| eGFR 30-60 mL/min/1.73 m<sup>2</sup> | 7 (54) |
| Kidney transplant recipient | 2 (15) |
| Unsure | 1 (8) |
| **Time with kidney disease (years)**<sup>†</sup> |  |
| Less than 5 | 3 (23) |
| 10-20 | 4 (31) |
| More than 20 | 2 (15) |
| Unsure or not applicable | 4 (31) |

\*Some participants selected more than one option.
<sup>†</sup>For patient participants or the corresponding patient with kidney disease for caregiver participants eGFR, estimated glomerular filtration rate

**Table 2.** Preferences within each topic area and corresponding prioritization results.

| Preferences discussed within each topic area | Vote Count | Priority Rank* |
| --- | --- | --- |
| **RECRUITMENT AND CONSENT PROCESS** |  |  |
| Technology use for recruitment by healthcare team | 11 | High |
| Consent provided by family member | 8 | High |
| Multiple methods for consent (e.g., one-on-one discussion, visual materials [posters, videos, etc.]) | 7 | Medium |
| Informed consent with knowledgeable and trusted person | 6 | Medium |
| Technology use for recruitment by research team | 4 | Low |
| Informed consent provided by responsible physician | 4 | Low |
| **INTERVENTION DELIVERY** |  |  |
| Acceptability of a new intravenous cannula if needed | 10 | High |
| Support and reimbursement for return visits to receive intervention | 9 | High |
| Intervention duration of more than 1 week | 7 | Medium |
| Intervention administration only during hospital admission | 5 | Medium |
| Acceptability of return visits after discharge to receive additional intervention doses if needed | 5 | Medium |
| Intervention duration of 1 week or less | 1 | Low |
| **TRIAL OUTCOMES** |  |  |
| Short- and long-term measures of kidney function (e.g., serum creatinine, need for dialysis, acute kidney injury severity) | 10 | High |
| Other patient health complications (e.g., cardiovascular events, death) | 8 | High |
| Healthcare utilization (e.g., hospital re-admission, emergency department visits, length of stay, nephrology follow-up) | 5 | Medium |
| Mental health outcomes (e.g., anxiety, depression) | 5 | Medium |
| Drug-related adverse events | 4 | Low |
| Activities of daily living, independence, and functional status (e.g., return to work) | 4 | Low |
| Physical health, symptoms, and patient-reported outcomes | 2 | Low |
| Caregiver outcomes (e.g., caregiving burden, mental health) | 2 | Low |

\*Priority assignment based on number of votes (i.e., dots), defined as high ( $\geq 7$ dots), medium (3-6 dots), and low ( $< 3$ dots) priority.

**Table 3.**
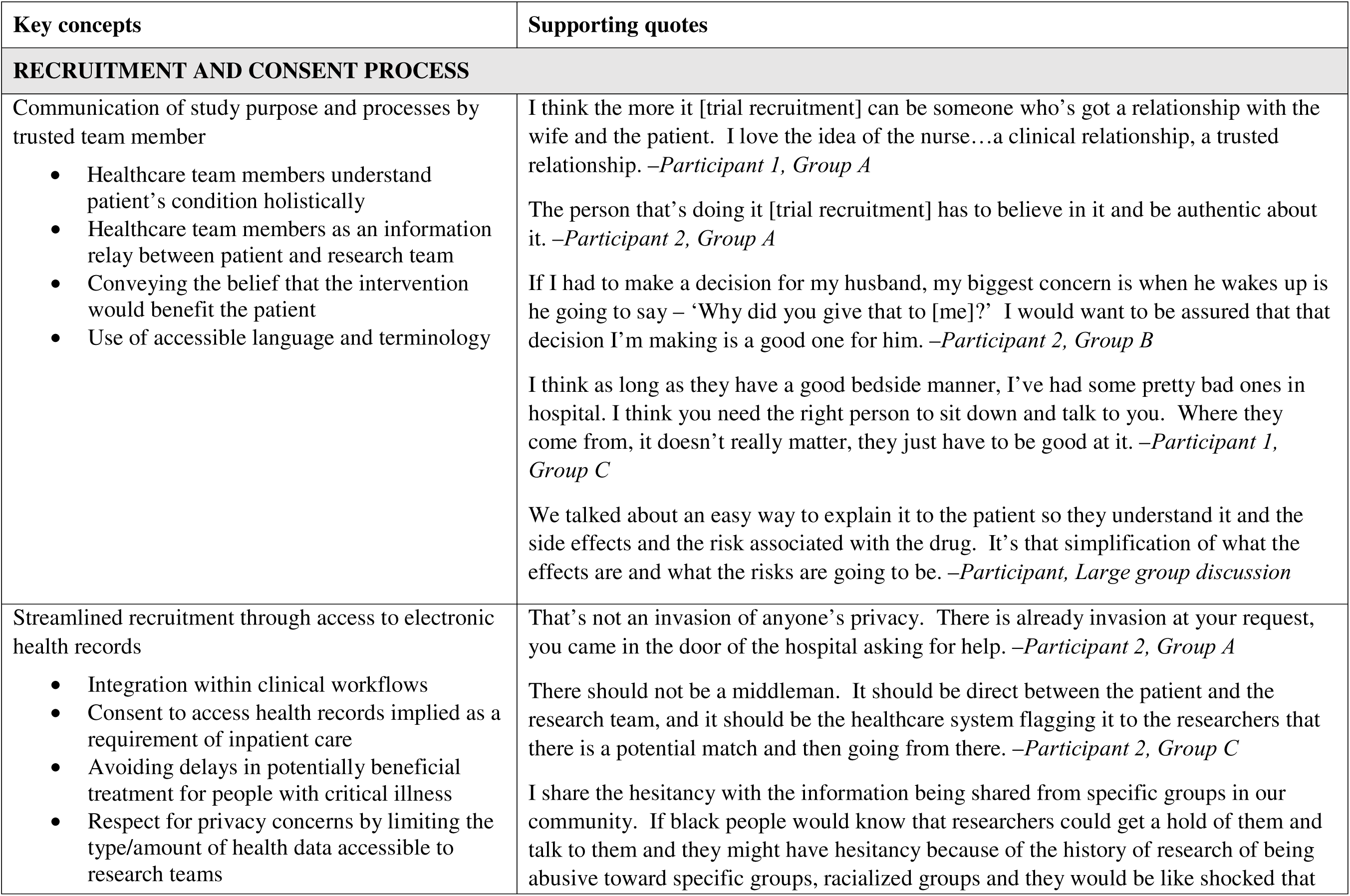
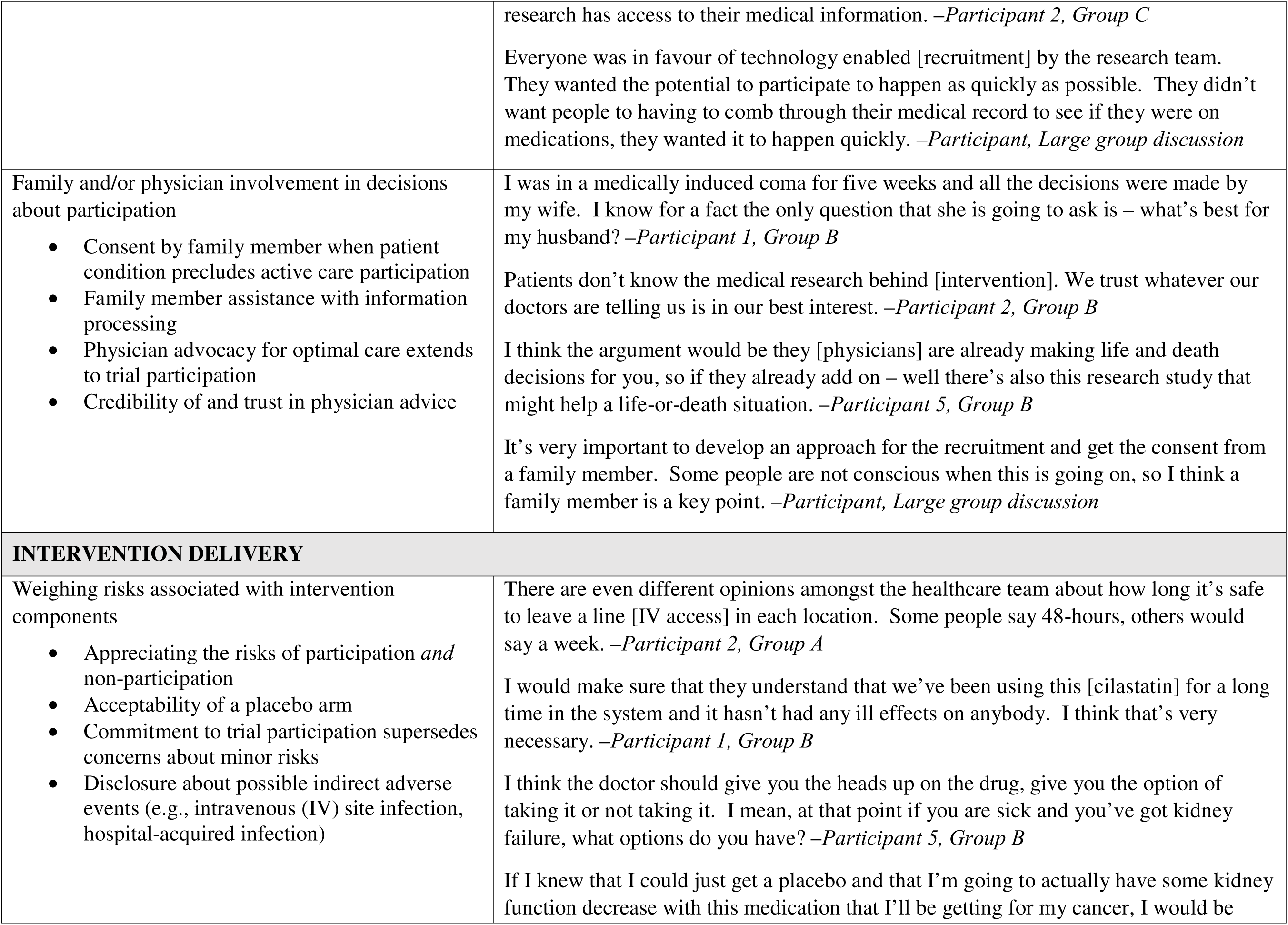
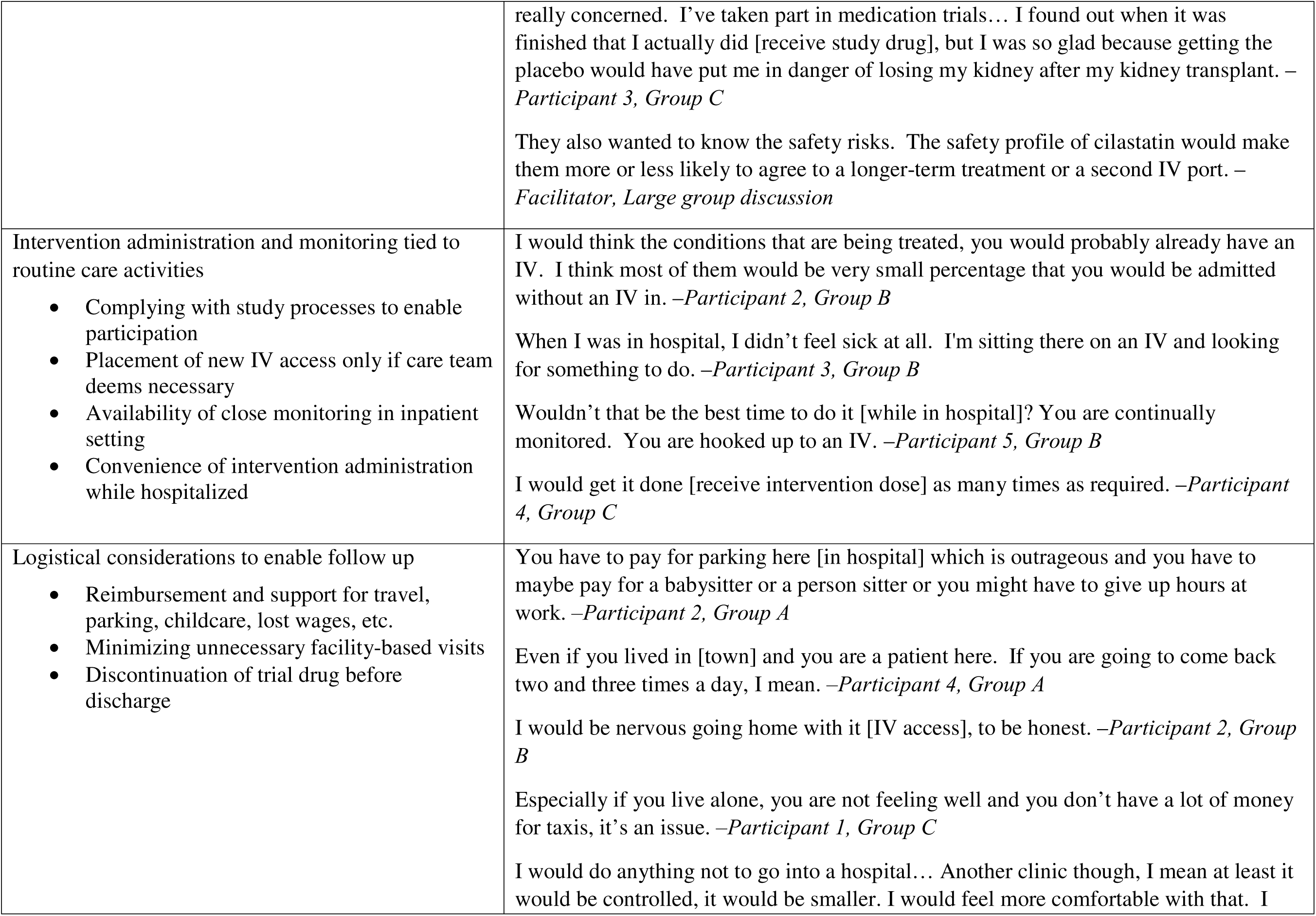
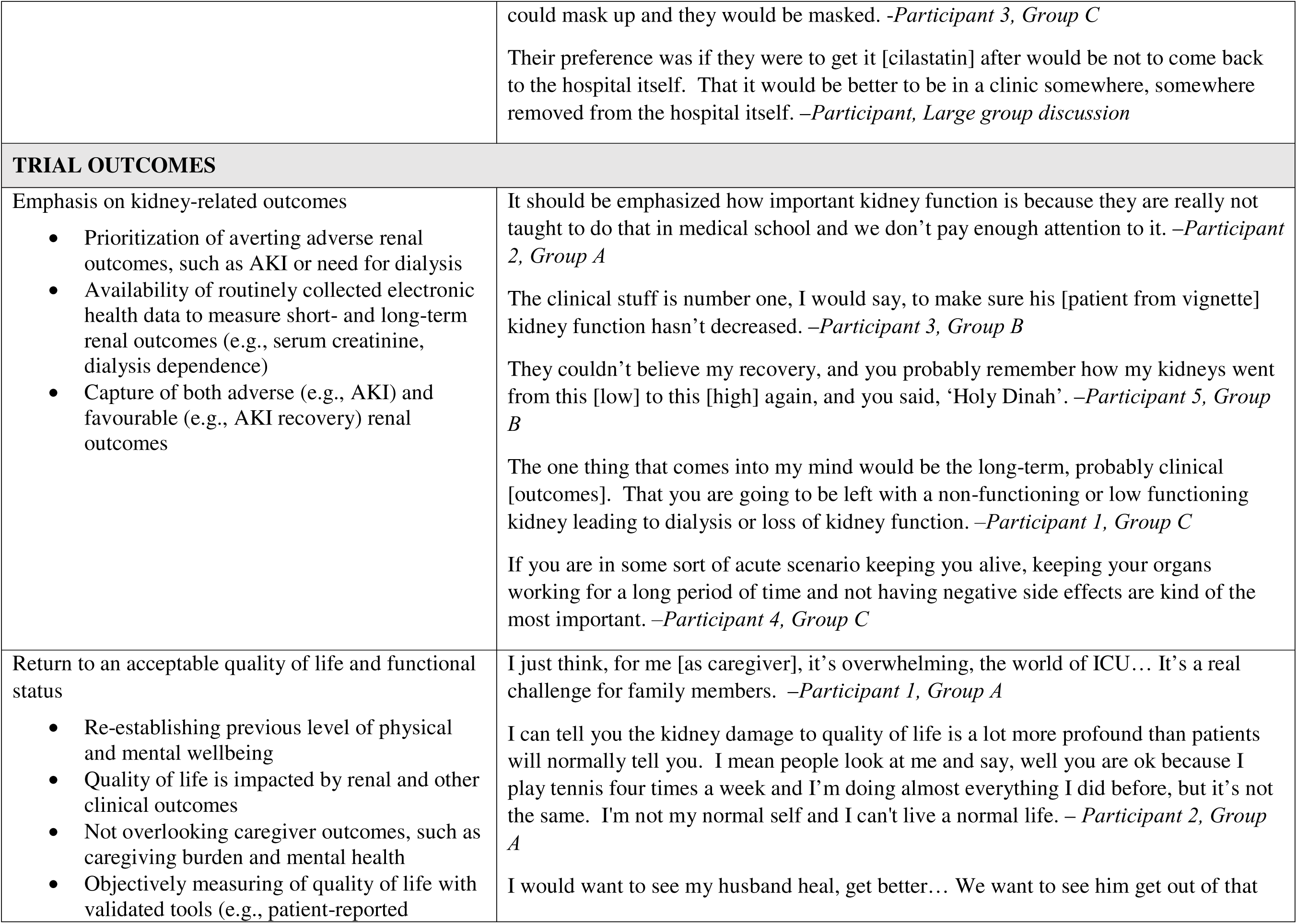
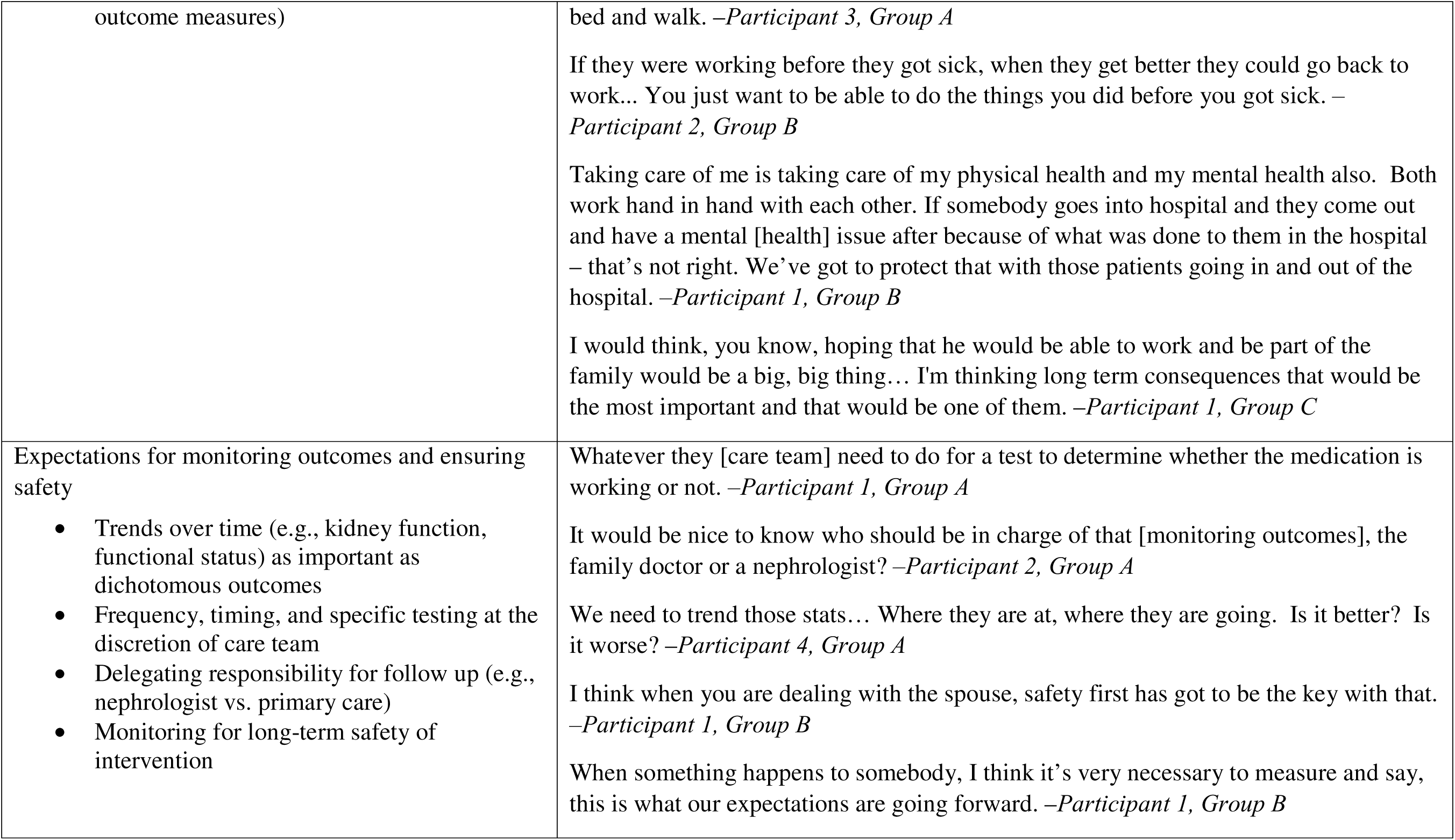
Thematic summary with key concepts and supporting quotes.

| Key concepts | Supporting quotes |
| --- | --- |
| **RECRUITMENT AND CONSENT PROCESS** |  |
| Communication of study purpose and processes by trusted team member<br>Healthcare team members understand patient's condition holistically<br>Healthcare team members as an information relay between patient and research team<br>Conveying the belief that the intervention would benefit the patient<br>Use of accessible language and terminology | I think the more it [trial recruitment] can be someone who's got a relationship with the wife and the patient. I love the idea of the nurse...a clinical relationship, a trusted relationship. *–Participant 1, Group A*<br>The person that's doing it [trial recruitment] has to believe in it and be authentic about it. *–Participant 2, Group A*<br>If I had to make a decision for my husband, my biggest concern is when he wakes up is he going to say – ‘Why did you give that to [me]?’ I would want to be assured that that decision I'm making is a good one for him. *–Participant 2, Group B*<br>I think as long as they have a good bedside manner, I've had some pretty bad ones in hospital. I think you need the right person to sit down and talk to you. Where they come from, it doesn't really matter, they just have to be good at it. *–Participant 1, Group C*<br>We talked about an easy way to explain it to the patient so they understand it and the side effects and the risk associated with the drug. It's that simplification of what the effects are and what the risks are going to be. *–Participant, Large group discussion* |
| Streamlined recruitment through access to electronic health records<br>Integration within clinical workflows<br>Consent to access health records implied as a requirement of inpatient care<br>Avoiding delays in potentially beneficial treatment for people with critical illness<br>Respect for privacy concerns by limiting the type/amount of health data accessible to research teams | That's not an invasion of anyone's privacy. There is already invasion at your request, you came in the door of the hospital asking for help. *–Participant 2, Group A*<br>There should not be a middleman. It should be direct between the patient and the research team, and it should be the healthcare system flagging it to the researchers that there is a potential match and then going from there. *–Participant 2, Group C*<br>I share the hesitancy with the information being shared from specific groups in our community. If black people would know that researchers could get a hold of them and talk to them and they might have hesitancy because of the history of research of being abusive toward specific groups, racialized groups and they would be like shocked that |
|  | research has access to their medical information. –*Participant 2, Group C*<br>Everyone was in favour of technology enabled [recruitment] by the research team. They wanted the potential to participate to happen as quickly as possible. They didn't want people to having to comb through their medical record to see if they were on medications, they wanted it to happen quickly. –*Participant, Large group discussion* |
| Family and/or physician involvement in decisions about participation<br>• Consent by family member when patient condition precludes active care participation<br>• Family member assistance with information processing<br>• Physician advocacy for optimal care extends to trial participation<br>• Credibility of and trust in physician advice | I was in a medically induced coma for five weeks and all the decisions were made by my wife. I know for a fact the only question that she is going to ask is – what's best for my husband? –*Participant 1, Group B*<br>Patients don't know the medical research behind [intervention]. We trust whatever our doctors are telling us is in our best interest. –*Participant 2, Group B*<br>I think the argument would be they [physicians] are already making life and death decisions for you, so if they already add on – well there's also this research study that might help a life-or-death situation. –*Participant 5, Group B*<br>It's very important to develop an approach for the recruitment and get the consent from a family member. Some people are not conscious when this is going on, so I think a family member is a key point. –*Participant, Large group discussion* |
| **INTERVENTION DELIVERY** |  |
| Weighing risks associated with intervention components<br>• Appreciating the risks of participation *and* non-participation<br>• Acceptability of a placebo arm<br>• Commitment to trial participation supersedes concerns about minor risks<br>• Disclosure about possible indirect adverse events (e.g., intravenous (IV) site infection, hospital-acquired infection) | There are even different opinions amongst the healthcare team about how long it's safe to leave a line [IV access] in each location. Some people say 48-hours, others would say a week. –*Participant 2, Group A*<br>I would make sure that they understand that we've been using this [cilastatin] for a long time in the system and it hasn't had any ill effects on anybody. I think that's very necessary. –*Participant 1, Group B*<br>I think the doctor should give you the heads up on the drug, give you the option of taking it or not taking it. I mean, at that point if you are sick and you've got kidney failure, what options do you have? –*Participant 5, Group B*<br>If I knew that I could just get a placebo and that I'm going to actually have some kidney function decrease with this medication that I'll be getting for my cancer, I would be |
|  | really concerned. I've taken part in medication trials... I found out when it was finished that I actually did [receive study drug], but I was so glad because getting the placebo would have put me in danger of losing my kidney after my kidney transplant. – *Participant 3, Group C*<br>They also wanted to know the safety risks. The safety profile of cilastatin would make them more or less likely to agree to a longer-term treatment or a second IV port. – *Facilitator, Large group discussion* |
| Intervention administration and monitoring tied to routine care activities<br>• Complying with study processes to enable participation<br>• Placement of new IV access only if care team deems necessary<br>• Availability of close monitoring in inpatient setting<br>• Convenience of intervention administration while hospitalized | I would think the conditions that are being treated, you would probably already have an IV. I think most of them would be very small percentage that you would be admitted without an IV in. – *Participant 2, Group B*<br>When I was in hospital, I didn't feel sick at all. I'm sitting there on an IV and looking for something to do. – *Participant 3, Group B*<br>Wouldn't that be the best time to do it [while in hospital]? You are continually monitored. You are hooked up to an IV. – *Participant 5, Group B*<br>I would get it done [receive intervention dose] as many times as required. – *Participant 4, Group C* |
| Logistical considerations to enable follow up<br>• Reimbursement and support for travel, parking, childcare, lost wages, etc.<br>• Minimizing unnecessary facility-based visits<br>• Discontinuation of trial drug before discharge | You have to pay for parking here [in hospital] which is outrageous and you have to maybe pay for a babysitter or a person sitter or you might have to give up hours at work. – *Participant 2, Group A*<br>Even if you lived in [town] and you are a patient here. If you are going to come back two and three times a day, I mean. – *Participant 4, Group A*<br>I would be nervous going home with it [IV access], to be honest. – *Participant 2, Group B*<br>Especially if you live alone, you are not feeling well and you don't have a lot of money for taxis, it's an issue. – *Participant 1, Group C*<br>I would do anything not to go into a hospital... Another clinic though, I mean at least it would be controlled, it would be smaller. I would feel more comfortable with that. I |
|  | could mask up and they would be masked. *–Participant 3, Group C*<br>Their preference was if they were to get it [cilastatin] after would be not to come back to the hospital itself. That it would be better to be in a clinic somewhere, somewhere removed from the hospital itself. *–Participant, Large group discussion* |
| **TRIAL OUTCOMES** |  |
| Emphasis on kidney-related outcomes<br>• Prioritization of averting adverse renal outcomes, such as AKI or need for dialysis<br>• Availability of routinely collected electronic health data to measure short- and long-term renal outcomes (e.g., serum creatinine, dialysis dependence)<br>• Capture of both adverse (e.g., AKI) and favourable (e.g., AKI recovery) renal outcomes | It should be emphasized how important kidney function is because they are really not taught to do that in medical school and we don't pay enough attention to it. *–Participant 2, Group A*<br>The clinical stuff is number one, I would say, to make sure his [patient from vignette] kidney function hasn't decreased. *–Participant 3, Group B*<br>They couldn't believe my recovery, and you probably remember how my kidneys went from this [low] to this [high] again, and you said, 'Holy Dinah'. *–Participant 5, Group B*<br>The one thing that comes into my mind would be the long-term, probably clinical [outcomes]. That you are going to be left with a non-functioning or low functioning kidney leading to dialysis or loss of kidney function. *–Participant 1, Group C*<br>If you are in some sort of acute scenario keeping you alive, keeping your organs working for a long period of time and not having negative side effects are kind of the most important. *–Participant 4, Group C* |
| Return to an acceptable quality of life and functional status<br>• Re-establishing previous level of physical and mental wellbeing<br>• Quality of life is impacted by renal and other clinical outcomes<br>• Not overlooking caregiver outcomes, such as caregiving burden and mental health<br>• Objectively measuring of quality of life with validated tools (e.g., patient-reported | I just think, for me [as caregiver], it's overwhelming, the world of ICU... It's a real challenge for family members. *–Participant 1, Group A*<br>I can tell you the kidney damage to quality of life is a lot more profound than patients will normally tell you. I mean people look at me and say, well you are ok because I play tennis four times a week and I'm doing almost everything I did before, but it's not the same. I'm not my normal self and I can't live a normal life. *– Participant 2, Group A*<br>I would want to see my husband heal, get better... We want to see him get out of that |
| outcome measures) | bed and walk. –*Participant 3, Group A*<br>If they were working before they got sick, when they get better they could go back to work... You just want to be able to do the things you did before you got sick. –<br>*Participant 2, Group B*<br>Taking care of me is taking care of my physical health and my mental health also. Both work hand in hand with each other. If somebody goes into hospital and they come out and have a mental [health] issue after because of what was done to them in the hospital – that’s not right. We’ve got to protect that with those patients going in and out of the hospital. –*Participant 1, Group B*<br>I would think, you know, hoping that he would be able to work and be part of the family would be a big, big thing... I’m thinking long term consequences that would be the most important and that would be one of them. –*Participant 1, Group C* |
| Expectations for monitoring outcomes and ensuring safety<br>• Trends over time (e.g., kidney function, functional status) as important as dichotomous outcomes<br>• Frequency, timing, and specific testing at the discretion of care team<br>• Delegating responsibility for follow up (e.g., nephrologist vs. primary care)<br>• Monitoring for long-term safety of intervention | Whatever they [care team] need to do for a test to determine whether the medication is working or not. –*Participant 1, Group A*<br>It would be nice to know who should be in charge of that [monitoring outcomes], the family doctor or a nephrologist? –*Participant 2, Group A*<br>We need to trend those stats... Where they are at, where they are going. Is it better? Is it worse? –*Participant 4, Group A*<br>I think when you are dealing with the spouse, safety first has got to be the key with that. –*Participant 1, Group B*<br>When something happens to somebody, I think it’s very necessary to measure and say, this is what our expectations are going forward. –*Participant 1, Group B* |

### Preferences for trial recruitment and consenting processes

Within the recruitment and consent topic area, participants highly prioritized healthcare team members’ access to electronic health records for identifying and recruiting eligible patients to the trial (11 votes). This included acceptance of a waiver of consent for access to health data to screen for eligibility based on potentially nephrotoxic medication exposures (i.e., technology enabled pre-screening). Participants also prioritized the ability of family members to provide informed consent for trial participation on behalf of patients who are critically ill and/or unable to provide consent themselves (8 votes).

During group discussions, participants emphasized the importance of rapport and “trusted relationships” with personnel approaching patients about trial participation, regardless of their role. Participants discussed wanting assurances from their care team that the intervention would be in their best interests and clear communication of the expectations of trial participation. Participants favoured the use of electronic health records for eligibility screening as a way of streamlining recruitment and avoiding delays in initiating a potentially life-saving therapy. While participants acknowledged privacy concerns, they suggested the benefits of prompt participant identification and recruitment through access to limited and necessary electronic health data outweighed these risks. Participants indicated that consent for trial participation should be provided by patients themselves with involvement of family members where possible, but that informed consent from family members or trusted physicians, who are responsible for “life and death decisions for you”, would be acceptable if patients’ condition precludes active care participation (e.g., sedated, critically ill, ventilated).

### Preferences for intervention delivery

In the prioritization exercise, most participants indicated that placement of a new intravenous access would be acceptable if needed to deliver the trial intervention (10 votes). Participants also identified the provision of support and reimbursement for return trial visits, whether for further intervention doses or for monitoring, as a high priority (9 votes).

Participants from all three small groups raised concerns about the acceptability of the trial’s placebo arm without prompting. Given the high stakes of AKI and potential benefits of the intervention, participants indicated it would be important to communicate to patients upfront that they have a 50% chance of receiving cilastatin and why a trial designed in this way is needed to establish its safety and efficacy. Participants prioritized their participation in a trial with the potential to avert AKI over details regarding precisely how and when cilastatin would be administered. However, they did indicate that use of an existing intravenous (IV) cannula would be preferrable to placing a new one, particularly for patients with difficult peripheral venous access or needle phobia, and that trial medication dosing should be coordinated with other routine care activities. Small group discussions also covered the convenience of intervention dosing and study monitoring as an inpatient, where patients are “continually monitored” and “hooked up to an IV”, and preferences for receiving the intervention drug only while hospitalized or, if required after discharge, through home visits. This related to participants’ expressed concerns about the safety of long-term intravenous cannulation and logistical challenges of having to return to hospital for ongoing treatments. They also relayed anticipated challenges to surveillance after hospital discharge, such as travel, parking costs, and lost wages, for which they expected the study team would provide support, as well as a preference to minimize facility-based study visits.

### Preferences for trial outcomes

Top priorities for trial outcomes included short- and long-term measures of kidney function (e.g., AKI, need for dialysis; 10 votes) and other clinical events (e.g., cardiovascular events, death; 8 votes).

Discussions about trial outcomes centered on averting adverse renal outcomes, specifically preventing AKI, AKI progression, and need for dialysis and leveraging routinely collected clinical and laboratory data for outcome ascertainment. Kidney and other clinical endpoints were largely discussed in relation to the complexity of hospitalized patients with AKI and the anticipated negative impact on quality of life and mental and physical health. Although quality of life outcomes were not prioritized during the voting exercise, participants discussed attaining one’s previous level of functional and mental wellbeing as an important long-term outcome.

They also identified a need to measure quality of life objectively using validated tools (e.g., patient-reported outcome measures) and in a way that considers the impact on both patients and caregivers. While they did not express a preference for specific instruments or tests to ascertain outcomes, participants valued trends in kidney function, quality of life, and functional status over time. Participants also raised concerns about the long-term safety of cilastatin and trial participation. They suggested that defining “expectations going forward” for monitoring over the course of the trial, including timing, responsible care team members, and long-term safety, would reassure participating patients and families and those considering enrolling in the trial.

### Evaluation survey

All participants (n=13) completed a post-workshop evaluation survey (Supplementary Table S3 and Figure S1) and indicated the workshop goals were communicated clearly and the materials were presented in an organized and well-paced way. Eleven participants stated that their opinions were captured in the large-group report-back summaries, and all but one participant felt the final voting results reflected the opinions and preferences discussed during the workshop. Three participants did not feel the vignettes added value to the workshop, with one suggesting they may have detracted from discussion about the experiences of the individual patient and caregiver participants.

## DISCUSSION

This consensus workshop was undertaken to explore the preferences and priorities of people with lived experience of AKI or risk factors for AKI, and to integrate these perspectives into the design of an upcoming trial for nephrotoxic AKI prevention. Over a series of group discussions and final voting exercise, participants identified priorities across aspects of trial design that included, most notably, the use of technology (i.e., access to electronic health records) and involvement of trusted individuals in trial recruitment, logistics of intervention administration (i.e., IV access, during hospitalization), support for study follow up, and emphasis on kidney-related and quality of life outcomes. While this workshop centered on the proposed trial intervention, cilastatin, our findings can also help other trials for AKI develop patient-centred approaches to recruitment and consent processes, intervention delivery, and outcome selection.

Low accrual rates and delays in identifying eligible patients put the viability of clinical trials at risk, and both are common when research staff must manually find participants for trials. Pre-screening potential participants for clinical trials can increase the efficiency of trial recruitment by quickly identifying people who may qualify for a study prior to approaching them and proceeding with informed consent. Technology enabled pre-screening is an increasingly accessible way to ensure systematic and timely identification of potential participants for modern-day trials as the availability of digital clinical information systems expands.^33^ Although privacy legislation governs how health data can be accessed in each jurisdiction, patient perspectives on use of such digital approaches are important to consider, as they may affect the willingness of patients to participate in trials. Technology enabled pre-screening is particularly relevant to trials seeking to enroll people with or at risk of AKI, as this population is widely distributed across hospital units and clinical services, making traditional manual approaches inefficient.

Participants identified the use of technology enabled pre-screening as a priority and considered access to this information by health care providers or members of the research team as acceptable. In fact, the use of algorithms within computer clinical information systems to identify people who would meet the eligibility criteria for a trial was the top ranked option for trial recruitment by our participants. Our findings are generally consistent with other research describing patient perspectives on the use of digital health tools in health research.^34–36^ In a recent literature review, Kassam et al. reported that most studies exploring this topic found that participants were willing to share personal health information digitally for clinical research provided there was clarity about who can access the information, for what purpose, and how privacy will be ensured.^34^ In a survey of the general population in the United States, Kim et al. found that participants were more willing to have their electronic health data shared when participants believed it would improve research quality and when they valued research benefit over privacy.^35^ A recent national survey exploring consent in the digital health ecosystem in Canada found that most respondents preferred data sources to be accessible by health care providers and delegates as the default option.^36^ These reports, in conjunction with our findings, imply a general patient acceptance and support for the use of technology enabled pre-screening approaches for clinical trials in AKI when the value of access to the information for the success of the trial is clear, and privacy and security concerns are appropriately addressed.

Participants in our workshop recognized the potential to avert nephrotoxic AKI through trial participation, which meant most, if not all, were willing to comply with processes for administering the intervention if recommended by research and care teams. However, they made suggestions for integrating intervention delivery with other routine care activities while in hospital to minimize burdens of clinical trial participation, such as need for additional testing and/or return visits, financial challenges, and concerns about safety.^37, 38^ Embedding trial processes within clinical workflows, such as timing cilastatin administration with other medications or coordinating laboratory tests with routine inpatient bloodwork, could further reduce the burden for healthcare team members and help to promote trial feasibility.^39^ Important concerns raised by participants about allocation to a placebo arm when the cilastatin intervention might help prevent poor AKI-associated outcomes align with patients’ perspectives on rare disease trial design from a qualitative study by Gaasterland et al.^40^ As patients view a novel intervention as a source of hope, the possibility of not receiving a potentially beneficial intervention may compromise this hope, the perceived benefits of trial participation, and ultimately patients’ willingness to enrol in trials of this design.^40^ In acknowledging these perspectives early in the design phase, recruitment materials and communication strategies can be co-developed with patients to clearly articulate trial processes and justification for a well-designed, placebo-controlled trial to establish intervention efficacy and safety.

Several narrative reviews have been published on the selection of outcomes for AKI, although these papers have focused on methodological aspects of outcome measures, limitations with the clinical criteria for ascertaining AKI, and requirements for regulatory drug approval.^41–46^ These existing publications have been exclusively written from the perspectives of clinical researchers, and although reports have called for greater patient participation in the study design of trials for AKI,^42^ few studies have explored patient priorities for AKI trial outcomes. The Standardised Outcomes in Nephrology (SONG) initiative is an international project that aims to establish core outcome measures across the spectrum of kidney disease for trials and other forms of research based on the shared priorities of patients, caregivers, clinicians, researchers, and policy makers.^47^ Although the SONG initiative has not addressed outcomes for AKI trials, findings from a focus group study conducted with patients and care providers of people with CKD may be relevant to trials for AKI, including the high priority assigned to outcomes of kidney function, mortality, fatigue, life participation, and mental health.^48^ We similarly found that patients prioritized measures of kidney function, both short and long term, as the top ranked trial outcome, followed by clinical outcomes including survival, cardiovascular events, and kidney failure for AKI trials. Notably, our findings from patients align with the most recent recommendations from AKI trialists, which highlight the occurrence of AKI as a key endpoint for phase 2B prevention or attenuation trials, and major adverse kidney events (including death, dialysis, or a sustained reduction in kidney function) for phase 3 AKI prevention, attenuation, or treatment trials.^41–43^

Our study is strengthened by the involvement of people with lived experience in workshop organization and the capture of diverse experiences and perspectives related to AKI. However, we acknowledge some limitations. First, the time allotted for small-group discussions may have been insufficient for participants to reflect and elaborate fully on important experiences, and some participants may have felt uncomfortable sharing their perspectives in this forum. We used skilled facilitators encouraged respectful interactions and ensure all participants had the opportunity to contribute. Second, while use of vignettes helped to focus small-group discussions on each of the topic areas, they may have unintentionally underemphasized participants’ own experiences. Third, the priorities brought forward to the voting exercise were compiled in real-time following small-group discussions, which means that preferences expressed by one participant or not discussed at length may not have been captured among voting options. However, results from the workshop evaluation survey suggest that participants felt the outcome reflected the content of group discussions. Fourth, the views and priorities of participants, who were largely white, cisgender, and highly educated, may not reflect those of underrepresented groups who are at risk of nephrotoxic AKI. Although priority categories are broad and discussions did cover aspects of socioeconomic disadvantage and health inequity, this area warrants future dedicated study. Lastly, the hybrid workshop format may have influenced the quality of interactions differentially between in-person and virtual attendees, although this drawback is outweighed by the inclusivity and diversity of participation enabled by the hybrid approach.

## CONCLUSION

In our consensus workshop, patients and caregivers prioritized technology enabled pre-screening and integration of trial processes and intervention delivery with routine care activities to streamline participation in a clinical trial of cilastatin for preventing nephrotoxic AKI. Participants’ prioritization of kidney-related and other clinical endpoints related in large part to their desire to avoid sequelae of AKI, such as dialysis dependence, and restore physical and mental wellbeing following hospitalization. The perspectives shared by patients and caregivers will uniquely inform development of our clinical trial protocol and can also help others to develop patient-centered approaches for recruitment and consent, intervention delivery, and outcome selection for AKI trials.

## DATA SHARING

All data produced in the present study are available upon reasonable request to the authors.

## DISCLOSURE

A pre-print version of this manuscript is available at MedRxivs: https://www.medrxiv.org/content/10.1101/2024.03.06.24303823v1

## Data Availability

All data produced in the present study are available upon reasonable request to the authors.

## REFERENCES

1. Steinman MA, Auerbach AD. Managing chronic disease in hospitalized patients. JAMA Intern Med. 2013;173(20): 1857–1858.

2. Mansur N, Weiss A, Beloosesky Y. Relationship of in-hospital medication modifications of elderly patients to postdischarge medications, adherence, and mortality. Ann Pharmacother. 2008;42(6): 783–789.

3. Krähenbühl-Melcher A, Schlienger R, Lampert M, Haschke M, Drewe J, Krähenbühl S. Drug-Related Problems in Hospitals. Drug Safety. 2007;30(5): 379–407.

4. Li L, Baker J, Rathnayake K, et al. Medication use and hospital-acquired acute kidney injury: an electronic health record-based study. Internal Medicine Journal. 2023;53(9): 1625–1633.

5. Perazella MA, Rosner MH. Drug-Induced Acute Kidney Injury. Clin J Am Soc Nephrol. 2022;17(8): 1220–1233.

6. James MT, Bhatt M, Pannu N, Tonelli M. Long-term outcomes of acute kidney injury and strategies for improved care. Nat Rev Nephrol. 2020;16(4): 193–205.

7. Tonelli M, Wiebe N, Guthrie B, et al. Comorbidity as a driver of adverse outcomes in people with chronic kidney disease. Kidney Int. 2015;88(4): 859–866.

8. James MT, Ghali WA, Knudtson ML, et al. Associations between acute kidney injury and cardiovascular and renal outcomes after coronary angiography. Circulation. 2011;123(4): 409–416.

9. James MT, Levey AS, Tonelli M, et al. Incidence and Prognosis of Acute Kidney Diseases and Disorders Using an Integrated Approach to Laboratory Measurements in a Universal Health Care System. JAMA Netw Open. 2019;2(4): e191795.

10. Collister D, Pannu N, Ye F, et al. Health Care Costs Associated with AKI. Clin J Am Soc Nephrol. 2017;12(11): 1733–1743.

11. Humanes B, Jado JC, Camaño S, et al. Protective Effects of Cilastatin against Vancomycin-Induced Nephrotoxicity. Biomed Res Int. 2015;2015: 704382.

12. Humanes B, Lazaro A, Camano S, et al. Cilastatin protects against cisplatin-induced nephrotoxicity without compromising its anticancer efficiency in rats. Kidney Int. 2012;82(6): 652–663.

13. Lau A, Chung H, Komada T, et al. Renal immune surveillance and dipeptidase-1 contribute to contrast-induced acute kidney injury. J Clin Invest. 2018;128(7): 2894–2913.

14. Hall AM, Trepiccione F, Unwin RJ. Drug toxicity in the proximal tubule: new models, methods and mechanisms. Pediatr Nephrol. 2022;37(5): 973–982.

15. Choudhury SR, Babes L, Rahn JJ, et al. Dipeptidase-1 Is an Adhesion Receptor for Neutrophil Recruitment in Lungs and Liver. Cell. 2019;178(5): 1205–1221.e1217.

16. Köller M, Brom J, Raulf M, König W. Cilastatin (MK 0791) is a potent and specific inhibitor of the renal leukotriene D4-dipeptidase. Biochem Biophys Res Commun. 1985;131(2): 974–979.

17. Hori Y, Aoki N, Kuwahara S, et al. Megalin Blockade with Cilastatin Suppresses Drug-Induced Nephrotoxicity. J Am Soc Nephrol. 2017;28(6): 1783–1791.

18. Tejedor A, Torres AM, Castilla M, Lazaro JA, de Lucas C, Caramelo C. Cilastatin protection against cyclosporin A-induced nephrotoxicity: clinical evidence. Curr Med Res Opin. 2007;23(3): 505–513.

19. Acharya D, Ghanim F, Harrison TG, et al. Nephroprotective effects of cilastatin in people at risk of acute kidney injury: A systematic review and meta-analysis. medRxiv. 2024: 2024.2003.2006.24303823.

20. Manafo E, Petermann L, Mason-Lai P, Vandall-Walker V. Patient engagement in Canada: a scoping review of the ‘how’ and ‘what’ of patient engagement in health research. Health Research Policy and Systems. 2018;16(1): 5.

21. Domecq JP, Prutsky G, Elraiyah T, et al. Patient engagement in research: a systematic review. BMC Health Services Research. 2014;14(1): 89.

22. Boote J, Baird W, Beecroft C. Public involvement at the design stage of primary health research: a narrative review of case examples. Health Policy. 2010;95(1): 10–23.

23. Skovlund PC, Nielsen BK, Thaysen HV, et al. The impact of patient involvement in research: a case study of the planning, conduct and dissemination of a clinical, controlled trial. Research Involvement and Engagement. 2020;6(1): 43.

24. Farah E, Kenney M, Kica A, Haddad P, Stewart DJ, Bradford JP. Beyond Participation: Evaluating the Role of Patients in Designing Oncology Clinical Trials. Curr Oncol. 2023;30(9): 8310–8327.

25. McMillan SS, King M, Tully MP. How to use the nominal group and Delphi techniques. Int J Clin Pharm. 2016;38(3): 655–662.

26. Mrklas KJ, Barber T, Campbell-Scherer D, et al. Co-Design in the Development of a Mobile Health App for the Management of Knee Osteoarthritis by Patients and Physicians: Qualitative Study. JMIR Mhealth Uhealth. 2020;8(7): e17893.

27. Dot Voting. 2018 Mar 02; https://www.ontario.ca/page/dot-voting.

28. Donald M, Beanlands H, Straus S, et al. Preferences for a self-management e-health tool for patients with chronic kidney disease: results of a patient-oriented consensus workshop. CMAJ Open. 2019;7(4): E713–E720.

29. Elliott MJ, Donald M, Farragher J, et al. Priorities for peer support delivery among adults living with chronic kidney disease: a patient-oriented consensus workshop. CMAJ Open. 2023;11(4): E736–E744.

30. Hsieh HF, Shannon SE. Three approaches to qualitative content analysis. Qual Health Res. 2005;15(9): 1277–1288.

31. Forero R, Nahidi S, De Costa J, et al. Application of four-dimension criteria to assess rigour of qualitative research in emergency medicine. BMC Health Serv Res. 2018;18(1): 120.

32. Staniszewska S, Brett J, Simera I, et al. GRIPP2 reporting checklists: tools to improve reporting of patient and public involvement in research. Res Involv Engagem. 2017;3: 13.

33. Aiyegbusi OL, Davies EH, Myles P, et al. Digitally enabled decentralised research: opportunities to improve the efficiency of clinical trials and observational studies. BMJ Evidence-Based Medicine. 2023;28(5): 328–331.

34. Kassam I, Ilkina D, Kemp J, Roble H, Carter-Langford A, Shen N. Patient Perspectives and Preferences for Consent in the Digital Health Context: State-of-the-art Literature Review. J Med Internet Res. 2023;25: e42507.

35. Kim KK, Sankar P, Wilson MD, Haynes SC. Factors affecting willingness to share electronic health data among California consumers. BMC Medical Ethics. 2017;18(1): 25.

36. Shen N, Kassam I, Zhao H, et al. Foundations for Meaningful Consent in Canada’s Digital Health Ecosystem: Retrospective Study. JMIR Med Inform. 2022;10(3): e30986.

37. Fogel DB. Factors associated with clinical trials that fail and opportunities for improving the likelihood of success: A review. Contemp Clin Trials Commun. 2018;11: 156–164.

38. Nicholls SG, Carroll K, Weijer C, et al. Ethical Issues in the Design and Conduct of Pragmatic Cluster Randomized Trials in Hemodialysis Care: An Interview Study With Key Stakeholders. Can J Kidney Health Dis. 2020;7: 2054358120964119.

39. Howard-Jones AR, Webb SA. Embedding clinical trials within routine health-care delivery: Challenges and opportunities. J Paediatr Child Health. 2021;57(4): 474–476.

40. Gaasterland CMW, van der Weide MCJ, du Prie-Olthof MJ, et al. The patient’s view on rare disease trial design - a qualitative study. Orphanet J Rare Dis. 2019;14(1): 31.

41. Legrand M, Bagshaw SM, Koyner JL, et al. Optimizing the Design and Analysis of Future AKI Trials. J Am Soc Nephrol. 2022;33(8): 1459–1470.

42. Zarbock A, Forni LG, Ostermann M, et al. Designing acute kidney injury clinical trials. Nature Reviews Nephrology. 2024;20(2): 137–146.

43. Lazzareschi D, Mehta RL, Dember LM, et al. Overcoming barriers in the design and implementation of clinical trials for acute kidney injury: a report from the 2020 Kidney Disease Clinical Trialists meeting. Nephrol Dial Transplant. 2023;38(4): 834–844.

44. Leaf DE, Waikar SS. End Points for Clinical Trials in Acute Kidney Injury. Am J Kidney Dis. 2017;69(1): 108–116.

45. Molitoris BA, Okusa MD, Palevsky PM, et al. Design of clinical trials in AKI: a report from an NIDDK workshop. Trials of patients with sepsis and in selected hospital settings. Clin J Am Soc Nephrol. 2012;7(5): 856–860.

46. Weisbord SD, Palevsky PM. Design of Clinical Trials in Acute Kidney Injury: Lessons from the Past and Future Directions. Semin Nephrol. 2016;36(1): 42–52.

47. Tong A, Manns B, Wang AYM, et al. Implementing core outcomes in kidney disease: report of the Standardized Outcomes in Nephrology (SONG) implementation workshop. Kidney Int. 2018;94(6): 1053–1068.

48. González AM, Gutman T, Lopez-Vargas P, et al. Patient and Caregiver Priorities for Outcomes in CKD: A Multinational Nominal Group Technique Study. American Journal of Kidney Diseases. 2020;76(5): 679–689.

