## Supplementary Material for "Patient preferences and priorities for the design of an acute kidney injury prevention trial: Findings from a consensus workshop"

**Supplemental Material**

**Supplementary Table S1.** Vignettes

**Supplementary Table S2.** Workshop topic guide

**Supplementary Table S3.** Post-workshop evaluation

**Supplementary Figure S1.** Summary of findings from post-workshop evaluation

**Supplementary Table S4.** Guidance for Reporting Involvement of Patients and Public (GRIPP2)

short form

**Supplementary Table S1.** Vignettes

**Vignette A - Edgar N. (patient)**

***Definitions:***

**Ramipril** – medication used to treat high blood pressure

**Metformin** – medication used to treat diabetes

**Atorvastatin** – medication used to treat high cholesterol

**Creatinine** – a blood test to measure kidney function

**eGFR** – a calculation based on creatinine that estimates how well the kidneys are filtering (i.e., what % they are working)

**Angiogram** – a test using contrast dye to determine if your coronary (heart) arteries are blocked

Edgar is a 74-year-old man with chronic kidney disease, high blood pressure, and diabetes. The medications he takes to treat these conditions include ramipril, metformin, and atorvastatin. He is a retired accountant and lives with his wife. He has two children and 6 grandchildren. He quit smoking about 10 years ago and drinks two glasses of wine per week.

Edgar is admitted to hospital with a heart attack, and his cardiologist tells him he needs to have a coronary angiogram to see if there are any blood vessel narrowings that could be treated with coronary stents or bypass surgery. His doctor tells him there is a chance his kidney function will get worse after the angiogram.

Edgar’s kidney function in hospital is similar to at home – creatinine is 150 umol/L, which is equivalent to 42% kidney function (i.e., eGFR 42 ml/min).

Edgar’s nurse mentions that he might be eligible for the PONTIAC trial.

**Vignette B – Elena R. (patient)**

Elena is a 32-year-old woman who was healthy and took no medications prior to her cervical cancer diagnosis 1 month ago. She has been in a relationship with her partner, Maria, for 6 years and has no children, although they have discussed plans for children in the future. She works as an elementary school teacher but has taken a leave of absence to focus on her health.

Together with her partner and cancer care team, Elena has decided on a treatment plan that includes radiation therapy and chemotherapy. She read on the internet that one of the chemotherapy medicines her care team plans on using, cisplatin, can cause kidney damage. She recalls her family doctor telling her that her kidneys were “normal” on recent bloodwork. Elena plans on asking her doctors about it when she comes into hospital for her cancer treatment next week.

Elena’s oncologist suggests she consider enrolling in the PONTIAC trial.

**Vignette C - Roberta D. (caregiver)**

***Definitions:***

**ICU** – Intensive Care Unit

**Sepsis** – a severe blood infection

**Metformin** – medication used to treat diabetes

**Ramipril** – medication used to treat high blood pressure

**Creatinine** – a blood test to measure kidney function

**eGFR** – a calculation based on creatinine that estimates how well the kidneys are filtering (i.e., what % they are working)

**Acute kidney injury** – sudden damage to the kidneys so that they cannot filter properly; potential causes include severe illness, certain medications, inflammation, or blockage of urine

Roberta is a 45-year-old woman who is married and has two school-aged children. Her husband, Michael, was admitted to ICU this morning for sepsis due to a bacteria called Staphylococcus aureus that they believe was caused by a skin infection on his leg. Michael struggles with obesity, has diabetes and sleep apnea, and takes metformin and ramipril. His kidney function was previously normal (above 60%, or eGFR greater than 60 ml/min), although he has had high amounts of albumin protein in his urine due to the diabetes.

When he presented to hospital, Michael had signs of acute kidney injury – his creatinine was elevated at 180 umol/L from his previous result of 80 umol/L 2 months ago. Because Michael is so sick, he requires medications to raise his blood pressure and a ventilator to help him breathe. Michael is sedated and cannot speak for himself, so his ICU care team speaks with Roberta about the proposed treatment plan that includes an antibiotic called vancomycin. They explain that in some people, this antibiotic can lead to further kidney injury.

The care team asks Roberta how she would like to proceed and mentions enrolment in the PONTIAC trial.

**Supplementary Table S2.** Workshop topic guide

Breakout session #1 – Topic: **Recruitment and consent processes**

| **Introduction** | In this session, we will be talking about options for the recruitment and consent processes for the trial. We want to explore your opinions on the different ways we could design the recruitment and consent processes because it can be challenging to identify eligible patients for clinical trials like this one and we want recruitment to be as efficient as possible (identify as many eligible patients as quickly as possible). This is one of the largest barriers to successfully completing clinical trials, so we would like to consider innovative approaches as long as they are felt to acceptable to patients. Let’s first spend a few minutes reviewing a vignette that we will use for this discussion.  **Vignette A – Edgar N. (patient)**  Edgar is a 74-year-old man with chronic kidney disease, high blood pressure, and diabetes. The medications he takes to treat these conditions include ramipril, metformin, and atorvastatin. He is a retired accountant and lives with his wife. He has two children and 6 grandchildren. He quit smoking about 10 years ago and drinks two glasses of wine per week.  Edgar is admitted to hospital with a heart attack, and his cardiologist tells him he needs to have a coronary angiogram to see if there are any blood vessel narrowings that could be treated with coronary stents or bypass surgery. His doctor tells him there is a chance his kidney function will get worse after the angiogram.  Edgar’s kidney function in hospital is similar to at home – creatinine is 150 umol/L, which is equivalent to 42% kidney function (i.e., eGFR 42 ml/min).  Edgar’s nurse mentions that he might be eligible for the PONTIAC trial. | |
| --- | --- | --- |
| **Questions/Discussion** | 1. What recruitment approach would you prefer? Why?   (e.g., Traditional, technology enabled through your health care provider, technology enabled to the research team)  Tell me more about how you feel about the acceptability of these approaches. (e.g. Balancing giving you the greatest opportunity to participate in the trial versus protecting your health care information and privacy)   2. How do you feel about a waiver of consent to access your health records to determine if you are eligible for the trial before you are approached for informed consent?   (e.g., using the information already captured in the hospital electronic medical record)   3. Tell me about how you or a member of your family (if you were too sick to make the decision for yourself) would want to be provided with information about the study as part of the informed consent process (eg. explanation from the study coordinator in person, a video explaining the study, via a telephone call with study coordinator, would you be comfortable receiving the information required to decide on participation from the doctor or nurse looking after you in hospital). | **Notes:** |

Breakout session #2 – Topic: **Intervention delivery**

| **Introduction** | In this group, we will be talking about the way the medical intervention, consisting of the drug cilastatin or placebo, would be delivered. We want to understand what you feel are acceptable ways to receive the medication so you would be willing to participate in the trial, recognizing that it must be delivered via an intravenous route (i.e. a needle into a blood vessel) and the time you are at risk of kidney damage from a drug could vary, and even extend beyond the time you are in hospital. Let’s spend a few minutes reviewing the vignette we will be using for this discussion.  **Vignette B – Elena R. (patient)**  Elena is a 32-year-old woman who was healthy and took no medications prior to her cervical cancer diagnosis 1 month ago. She has been in a relationship with her partner, Maria, for 6 years and has no children, although they have discussed plans for children in the future. She works as an elementary school teacher but has taken a leave of absence to focus on her health.  Together with her partner and cancer care team, Elena has decided on a treatment plan that includes radiation therapy and chemotherapy. She read on the internet that one of the chemotherapy medicines her care team plans on using, *cisplatin*, can cause kidney damage. She recalls her family doctor telling her that her kidneys were “normal” on recent bloodwork. Elena plans on asking her doctors about it when she comes into hospital for her first cancer treatment next week.  Elena’s oncologist suggests she consider enrolling in the PONTIAC trial. | |
| --- | --- | --- |
| **Questions/Discussion** | 1. How would you feel about participating in the trial with the intervention requiring a placement of an intravenous (IV) needle?   (if I already had an IV in place, if I didn’t already have an IV and had to have one placed)  Do you think this would influence your participation in the trial?   2. How would you feel about receiving the cilastatin or placebo at multiple times in hospital or clinic visits when you are being exposed to nephrotoxic medication and would this affect your participation in the trial? 3. Would knowing you would have to return to a clinic or receive intravenous medication after you are discharged home influence your willingness to participate in the trial? | **Notes:** |

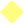

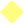

Breakout session #3 – Topic: **Patient-prioritized outcomes**

| **Introduction** | In this group, we will be talking about the selection of outcomes that will be measured to determine how effective the medication is. We want to identify the outcomes that are most important for patients. Clinical trials must identify a primary outcome, which will determine the number of patients required to be enrolled in the study to be confident in the results. Other outcomes can also measured and should be pre-specified before the study starts, though the additional costs and complexity of measuring them are important considerations.  **Vignette C – Roberta D. (caregiver)**  Roberta is a 45-year-old woman who is married and has two school-aged children. Her husband, Michael, was admitted to ICU this morning for sepsis due to a bacteria called Staphylococcus aureus that they believe was caused by a skin infection on his leg. Michael struggles with obesity, has diabetes and sleep apnea, and takes metformin and ramipril. His kidney function was previously normal (above 60%, or eGFR greater than 60 ml/min), although he has had high amounts of albumin protein in his urine due to the diabetes.  When he presented to hospital, Michael had signs of acute kidney injury – his creatinine was elevated at 180 umol/L from his previous result of 80 umol/L 2 months ago. Because Michael is so sick, he requires medications to raise his blood pressure and a ventilator to help him breathe. Michael is sedated and cannot speak for himself, so his ICU care team speaks with Roberta about the proposed treatment plan that includes an antibiotic called vancomycin. They explain that in some people, this antibiotic can lead to further kidney injury.  The care team asks Roberta how she would like to proceed and mentions enrolment in the PONTIAC trial. | |
| --- | --- | --- |
| **Questions/Discussion** | 1. Review the list and description of the different categories of clinical outcomes (these include surrogate and patient-important clinical outcomes, patient-reported health status measures, and measures of experience)? Which of these are important outcomes to you? 2. Which outcomes do you feel are most important to you? 3. Which type of patient reported health status measure would you want to know whether the intervention could improve? | **Notes:** |

**Supplementary Table S3.** Post-workshop evaluation

|  | Strongly  disagree |  |  |  | Strongly  agree |
| --- | --- | --- | --- | --- | --- |
|  | 1 | 2 | 3 | 4 | 5 |
| 1. The goal of the workshop was described clearly. | ❑ | ❑ | ❑ | ❑ | ❑ |
| 2. The program was well paced within the allotted time. | ❑ | ❑ | ❑ | ❑ | ❑ |
| 3. The facilitators were good communicators. | ❑ | ❑ | ❑ | ❑ | ❑ |
| 4. The material was presented in an organized manner. | ❑ | ❑ | ❑ | ❑ | ❑ |
| 5. The vignettes aided the topic discussion. | ❑ | ❑ | ❑ | ❑ | ❑ |
| 6. The facilitators were knowledgeable about the topic. | ❑ | ❑ | ❑ | ❑ | ❑ |
| 7. My opinions were captured in the summaries provided by the facilitators back to the large group and in the voting options listed at the end of the workshop. | ❑ | ❑ | ❑ | ❑ | ❑ |
| 8. I feel the final voting results reflect the opinions and discussions of patients who participated in the workshop. | ❑ | ❑ | ❑ | ❑ | ❑ |
| 9. Given the objectives, this workshop was: | ❑ Too short ❑ Right length ❑ Too long | | | | |
| 10. Please rate the following: | Excellent | Very good | Good | Fair | Poor |
| a. Workshop reading material | ❑ | ❑ | ❑ | ❑ | ❑ |
| b. Workshop organization | ❑ | ❑ | ❑ | ❑ | ❑ |
| c. Instructions | ❑ | ❑ | ❑ | ❑ | ❑ |
| What did you most appreciate/enjoy/think was best about the workshop?  Do you have any suggestions for improvement? | | | | | |

**Supplementary Figure S1.** Summary of findings from post-workshop evaluation

Twelve of the thirteen participants completed the evaluation. The following graph illustrates participant responses to the evaluation questions:

Total Number of Responses (N=12)

| Opinions About the Workshop | Strongly Disagree | Somewhat Disagree | Neither agree nor disagree | Somewhat agree | Strongly agree |
| --- | --- | --- | --- | --- | --- |
| The goal of the workshop was described clearly | 0 | 0 | 0 | 1 | 11 |
| The workshop was well paced within the allotted time. | 0 | 0 | 0 | 5 | 6 |
| The facilitators were good communicators. | 0 | 0 | 1 | 1 | 10 |
| The material was presented in an organized manner. | 0 | 0 | 0 | 2 | 9 |
| The vignettes aided the topic discussion. | 1 | 0 | 2 | 6 | 3 |
| The facilitators were knowledgeable about the topic | 1 | 0 | 0 | 1 | 10 |
| My opinions were captured in the summaries provided by the facilitators back to the large group and in the voting options listed at the end of the workshop | 0 | 1 | 1 | 3 | 7 |
| I feel the final voting results reflect the opinions and discussions of patients who participated in the workshop | 0 | 1 | 0 | 1 | 10 |

All participants agreed that the goal of the workshop was clearly described. Nine participants either strongly or somewhat agreed that the vignettes aided the discussion around the three topic areas. Most participants reported that the final voting results accurately reflected the discussion and their opinions (11/12), while 7/12 participants felt that their opinions were captured in summaries presented by the facilitators during the large group sessions.

**Supplementary Table S4.** Guidance for Reporting Involvement of Patients and Public (GRIPP2) short form

| **Section and Topic** | **Item** | **Reported on page No** |
| --- | --- | --- |
| 1. Aim | Report the aim of PPI in the study | 5 |
| 2. Methods | Provide a clear description of the methods used for PPI in the study | 5-9 |
| 3. Study results | Outcomes-Report the results of PPI in the study, including both positive and negative outcomes | 9-13 |
| 4. Discussion and conclusions | Outcomes-Comment on the extent to which PPIs influenced the study overall. Describe positive and negative effects | 13-17 |
| 5. Reflections/critical perspective | Comment critically on the study, reflecting on the things that went well and those that did not, so others can learn from this experience | 16-17 |

PPI=patient and public involvement
